# Barriers and Facilitators to Advance Care Planning Implementation for Patients with Neurodegenerative Diseases among Indian Physicians: A Mixed-Methods Analysis

**DOI:** 10.1101/2025.10.17.25338242

**Authors:** Pavit Singh, Parvathy, Deepa Dash, Nishkarsh Gupta, Usha Ramanathan, Teneille E. Gofton, Claudia Chou, Soaham Desai, Pramod Kumar Pal, Roopkumar Gursahani, Baikuntha Panigrahi, Banusri Velpandian, Prasun Chatterjee, Avinash Chakrawarty, Anup Singh, Suman Kushwaha, K.P. Divya, Lakshmi Narasimhan Ranganathan, Manjari Tripathi, Deepti Vibha, Rajesh Kumar Singh, Animesh Das, Jasmine Parihar, Sumit Malhotra, Ashish Dutt Upadhyay, Abhishek Pathak, Arunmozhimaran Elavarasi

## Abstract

**Background:** Advance care planning (ACP) is a process that enables individuals to define and communicate their goals and preferences for future medical care, which is especially important in chronic progressive illnesses, such as Parkinson’s disease (PD) and other neurodegenerative disorders. Despite its recognized benefits in improving patient autonomy and end-of-life care outcomes, ACP remains underutilized in India. This study aimed to assess the attitudes and practices of Indian neurologists and geriatricians regarding ACP, identify perceived barriers, and suggest strategies to improve uptake.

**Methods:** A mixed-methods approach was employed in this study. In the first phase, a structured online survey was distributed to physicians across India who cared for patients with PD and neurodegenerative disorders. The survey collected demographic details and ACP-related practices, attitudes, and perceived barriers to ACP. In the second phase, in-depth qualitative interviews were conducted with a purposively sampled subset of respondents, and inductive thematic analysis was performed to gain deeper insights.

**Results:** A total of 140 physicians participated in this survey. Although 93.6% acknowledged the necessity of ACP, only 25% felt that they had sufficient time, and 20% perceived that they had adequate resources for meaningful discussions. Lack of legal clarity (52.1%), training (16.4%), and institutional support (65.7%) were commonly cited as barriers. Qualitative interviews with 15 respondents revealed additional challenges, such as concerns about provoking hopelessness and denial from patients and families. The interviews also revealed that physicians confused ACP with advanced treatment strategies or treatment of advanced disease. Nevertheless, some physicians shared positive experiences, noting that early personalized discussions improved trust and communication, which could facilitate uptake of ACP.

**Conclusions:** We identified several systemic, professional, and physician-perceived sociocultural barriers that hinder ACP implementation. To bridge this gap, legal reforms, structured ACP training, and public awareness initiatives are necessary. Tailored culturally sensitive models involving multidisciplinary teams may improve ACP adoption within the Indian context. The term’ advance care planning’ may need to be replaced by other terminology, such as ‘Future care planning,’ which may avoid confusion with other domains of care in Parkinson’s disease and other Neurodegenerative conditions.

## Introduction

Advance care planning (ACP) is a structured process through which individuals articulate their goals, values, and preferences for future medical treatment and care. This involves engaging in conversations with family and healthcare providers, and documenting and revisiting these choices when appropriate(1). It is an important process that empowers patients and their care partners by providing opportunities to consider and share their beliefs, preferences, and wishes about decisions regarding future medical care while they retain decision-making capacity (2).

Evidence suggests that ACP improves the quality of patient-clinician communication, reduces unwanted hospitalizations, increases the use of palliative care (3), enhances patients’ quality of life and autonomy, and helps them to die with comfort (3). Because ACP is an essential domain for providing high-quality end-of-life care, all physicians should be encouraged to openly discuss end-of-life care at appropriate times, and discuss and determine the preferences and wishes of the patient, for their care throughout the disease course, based on their understanding of the patient’s disease course (4,5).

Despite documented benefits in the Western world, ACP uptake is limited in India. In an Internet survey of graduate Indians by Dhru et al. (6), only 28% reported having some knowledge of AMDs.

In India, the concept of an Advance Medical Directive (AMD)—also referred to as a living will, wherein a competent adult, while retaining decision-making capacity, could make directives about future medical treatment—was legally recognized by the Supreme Court in its landmark 2018 *Common Cause(*A Registered Society) *vs. Union of India* judgment. The Court affirmed that the right to die with dignity is part of the fundamental right to life under Article 21 and permitted the use of AMDs to guide end-of-life decisions, including the withdrawal or withholding of life-sustaining treatment in terminally ill patients. However, the 2018 guidelines involved complex procedures, such as judicial magistrate attestation and multiple medical board reviews, which limited practical implementation. In January 2023, the Supreme Court of India issued a clarificatory order in the aforementioned case, refining and simplifying the procedures for executing AMDs.(7) This ruling marked a significant stride forward in enabling Advance Care Planning (ACP) in India. By replacing the onerous requirement of a Judicial Magistrate of First Class (JMFC) countersignature with more practical attestation by a notary or gazetted officer, and streamlining how these directives are recorded, preserved, and acted upon, the Court significantly enhanced its accessibility and feasibility. Noticing the lack of awareness related to ACP, in 2025, a collective of healthcare professionals, legal experts, and community advocates launched the Advance Care Planning India (ACP India) to fill the gaps.

ACPs’ role in improving the quality of death of people living with Parkinson’s disease and other neurodegenerative conditions has not been explored so far in India. ACP is an essential component in the management of Parkinson’s disease, due to its complex, progressive course, and high burden of complications such as dementia, infections, and falls. Importantly, the symptom burden in late-stage PD is comparable to that of end-stage cancer. (8) Considering the relevance of this ACP, it becomes necessary to understand the barriers and facilitators associated with the update of ACP in India to design institutional and health system-level policies to promote its uptake. The optimal timing and structure for ACP in PD and other neurodegenerative conditions, combined with reported gaps in healthcare providers’ confidence and competence in delivering ACP, underscores the need to better understand and support ACP in this population. (9,10)

In this mixed-methods study, we aimed to analyze the attitudes and practices of physicians treating patients with Parkinson’s disease and other neurodegenerative diseases in India, as well as to understand the barriers and facilitators to the implementation of ACP.

## Materials and Methods

This study employed a sequential mixed-methods design comprising two parts. First, we conducted a cross-sectional survey of physicians, using both multiple-choice and open-ended questions, to assess their attitudes, self-reported practices, and approaches to ACP. The qualitative component involved in-depth interviews with a subset of respondents selected through purposive sampling to gain deeper insights into their perspectives and experiences with ACP. Rigor was ensured through triangulation at multiple levels. Data triangulation was achieved by comparing the survey findings with in-depth interviews. Methodological triangulation combines quantitative measures with qualitative exploration, allowing convergence and complementarity of the results. Investigator triangulation was maintained through reflexive team discussion. Analytical triangulation was performed by integrating both datasets under the Consolidated Framework for Implementation Research (CFIR) framework, highlighting convergent, complementary, and divergent findings.

### 2.1 Qualitative Study

#### 2.1.1 Survey Design

Based on a thorough literature review, we designed a survey to address gaps in the current knowledge. The study was designed according to the CHERRIES checklist for online surveys (11). Survey questions were developed by two authors (PS and PK). The face validity of the questionnaire was evaluated by neurologists (AE and DD) and a palliative care specialist (NG) to assess the wording, structure, layout, and readability and to provide an estimate of the time required to complete the questions. The questionnaire was revised and adapted accordingly, and content validity was assessed qualitatively and iteratively (NG, AE, and DD) to ensure coverage of all intended domains. The Grammarly software (Grammarly Inc., San Francisco, CA, USA) ensured the readability, correctness, and engagement of the survey.

The questionnaire consisted of two sections (see Tables 1 and 2). All questions were mandatory for the survey to be completed and submitted, and the completeness check feature of Google Forms (Google LLC, Mountain View, CA, USA) ensured this. The first section pertained to the demographics, practice setting, and number of patients with Parkinson’s disease and other neurodegenerative disorders seen per week. The second section contained 15 multiple-choice questions regarding ACP awareness and practices. We asked two open-ended questions regarding ACP experiences. The average time to complete the survey was 5 minutes. The study and survey questionnaires were approved by the Institutional Review Board.

**Table 1.** Demographic details.

| Parameter | Total Responses<br>n(%) | Those who underwent in-depth interviews (15) | Those who did not undergo IDI (125) | P value |
| --- | --- | --- | --- | --- |
| 1. Age group of respondents |  |  |  | 0.12 |
| a) 20-30 years | 44(31.4) | 2(13.33) | 42(33.6) |  |
| b) 31-40 years | 62(44.3) | 6(40.0) | 56(44.8) |  |
| c) 41-50 years | 17(12.1) | 3(20.0) | 14(11.2) |  |
| d) 51-60 years | 12(8.6) | 3(20.0) | 9(7.2) |  |
| e) Above 60 | 5(3.6) | 1(6.7) | 4(3.2) |  |
| 2. Average number of persons living with Parkinson's disease and other neurodegenerative diseases consulted per week |  |  |  | 0.67 |
| a) Less than 10 | 68(48.6) | 6(40.0) | 62(49.6) |  |
| b) Between 10 and 50 | 64(45.7) | 8(53.3) | 56(44.8) |  |
| c) More than 50 | 8(5.7) | 1(6.7) | 7(5.6) |  |
| 3. Practice setting |  |  |  | 0.29 |
| a) Government Medical college/Teaching institute | 86(61.4) | 6(40.0) | 80(64.0) |  |
| b) Tertiary care, Private hospital | 34(24.3) | 3(20.0) | 31(24.8) |  |
| c) Private Clinic | 20(14.3) | 6(40.0) | 14(11.2) |  |
| 4. Level of training |  |  |  | 0.002 |
| a) Neurologist | 60(42.9) | 13(86.7) | 47(37.6) |  |
| b) Neurology Resident | 45(32.1) | 1(6.7) | 44(35.2) |  |
| c) Specialist training in movement disorders | 11(7.9) | 1(6.7) | 10(8.0) |  |
| d) General physician/Geriatrician | 24(17.1) | 0(0.0) | 24(19.2) |  |
| 5. Exposure to training in advance care planning |  |  |  | 0.80 |
| a) Formal training during residency/fellowship | 59(42.1) | 8(53.3) | 51(40.8) |  |
| b) Short-course training or workshops in medical education | 14(10.0) | 1(6.7) | 13(10.4) |  |
| c) Self-initiated training through online modules | 9(6.4) | 1(6.7) | 8(6.4) |  |
| d) Informal exposure through clinical experience | 35(25.0) | 3(20.0) | 32(25.6) |  |
| e) No exposure | 23(16.4) | 2(13.3) | 21(16.8) |  |
| 6. Participants willing to give an interview |  |  |  |  |
| a) Yes | 50(35.7) |  |  |  |
| b) No | 90(64.3) |  |  |  |

**Table 2.** Attitude of physicians towards advanced care planning.

| Parameter | Responses<br>(Percentage) | Those who<br>provided IDI | Those who<br>did not<br>provide IDI | P value |
| --- | --- | --- | --- | --- |
| **Necessity of discussing ACP** |  | N=15 | N=125 | 0.25 |
| a) Yes | 131(93.6) | 13(86.7) | 118(94.4) |  |
| b) Depends on the patient initiating the conversation - opt in | 9(6.4) | 2(13.3) | 7(5.6) |  |
| c) No | 0 | 0 | 0 |  |
| **Time to introduce ACP discussion** |  |  |  | 0.37 |
| a) At the time of diagnosis | 72(51.4) | 5(33.3) | 67(53.6) |  |
| b) At the time of the onset of motor fluctuations/cognitive deterioration | 30(21.4) | 5(33.33) | 25(20.0) |  |
| c) At the time of needing support for activities of daily living | 34(24.3) | 5(33.3) | 29(23.2) |  |
| d) When non-motor symptom worsens/the patient needs hospitalization. | 4(2.9) | 0(0.0) | 4(3.2) |  |
| **Person to initiate conversation regarding ACP** |  |  |  | 0.48 |
| a) By the Primary treating neurologist | 76(54.3) | 9(60.0) | 67(53.6) |  |
| b) By trained nurse practitioners/patient care managers in the clinic | 23(16.4) | 4(26.7) | 19(15.2) |  |
| c) By physicians with specialist palliative care training | 33(23.6) | 2(13.3) | 31(24.8) |  |
| d) By pamphlets provided in the physicians' clinic | 0 | 0(0.0) | 0(0.0) |  |
| e) By support groups | 8(5.7) | 0(0.0) | 8(6.4) |  |
| **Time sufficiency to discuss ACP** |  |  |  | 0.59 |
| a) Mostly | 35(25) | 2(13.3) | 33(26.4) |  |
| b) Sometimes | 90(64.3) | 11(73.3) | 79(63.2) |  |
| c) Never | 15(10.7) | 2(13.3) | 13(10.4) |  |
| **Adequate Resources to discuss ACP** |  |  |  | 0.55 |
| a) Yes | 28(20) | 2(13.3) | 26(20.8) |  |
| b) No | 92(65.7) | 12(80.0) | 80(64.0) |  |
| c) Not sure | 20(14.3) | 1(6.7) | 19(15.2) |  |
| **Comfort in discussing ACP** |  |  |  | 0.38 |
| a) Yes, always | 54(38.6) | 8(53.3) | 46(36.8) |  |
| b) Mostly | 50(35.7) | 6(40.0) | 44(35.2) |  |
| c) Sometimes | 31(22.1) | 1(6.7) | 30(24.0) |  |
| d) No, I am not comfortable | 5(3.6) | 0(0.0) | 5(4.0) |  |
| **Opinions regarding ACP** |  |  |  |  |
| a) I am not aware of Advance Health directives. | 22(15.7) | 1(6.7) | 21(16.8) | 0.47 |
| b) I feel that I am inexperienced in holding this conversation | 32(22.9) | 3(20.0) | 29(23.2) | >0.99 |
| c) I feel I need more information on the legal aspects of Advance care planning in India | 73(52.1) | 6(40.0) | 67(53.6) | 0.32 |
| d) I feel that patients may not be interested in ACP in the Indian setting, as they may not value planning for future medical situations or understand prognosis | 35(25.0) | 6(40.0) | 29(23.2) | 0.16 |
| e) I believe it will make the patient feel abandoned and cause family distress | 15(10.7) | 3(20.0) | 12(9.6) | 0.20 |
| f) I feel apprehensive that patients/families will respond inappropriately | 17(12.14) | 1(6.7) | 16(12.8) | 0.70 |
| g) I feel that discussing advanced care planning is equivalent to giving up on the patient and abandoning the patient | 2(1.4) | 0(0.0) | 2(1.6) | >0.99 |
| h) I feel that it is not rewarding for me as a physician who is trained to diagnose and treat patients, and not discussing values and preferences for future medical situations | 3(2.1) | 0(0.0) | 3(2.4) | >0.99 |
| i) I feel that initiating this discussion with patients will increase trust and improve the quality-of-life outcomes in patients and families | 86(61.4) | 12(80.0) | 74(59.2) | 0.16 |

#### 2.1.2 Survey dissemination

We sent an open survey using a self-administered questionnaire via Google Forms. Before distributing the questionnaire, the survey was thoroughly pre-tested for ease of use and technical functionality in four individuals who were neither part of the study team nor respondents to the survey. The questionnaire was circulated by one author (initials here) in closed professional groups on social media platforms such as WhatsApp and email groups of academic institutions that agreed to circulate, with an estimated reach of around 1000 physicians, including academic and community practitioners of Neurology and geriatric medicine, and neurologists specializing in movement disorders in India from October 2024 to March 2025. However, the exact reach of the survey is unknown, since the survey was not advertised and the participants did not receive any incentive to respond. The survey forms indicated that the survey would be de-identified for analysis and would be completely confidential and anonymous to avoid social desirability bias, thereby encouraging participants to be more comfortable and honest when answering questions (12).

Participation in the survey was entirely voluntary. The respondents were informed of the purpose of the study and the expected duration of the survey, and they provided their consent before beginning the survey. We ensured that no duplicate responses were submitted by limiting each participant to one survey response using Google Forms. Survey respondents could edit their responses during submission, but they could not access the survey or change responses afterwards. The login details were stored along with the survey results and subsequently removed during the de-identification process. The results were stored in secure, encrypted storage systems prior to analysis.

### 2.2 Qualitative Study

#### 2.2.1 Interview design

We conducted in-depth interviews with a subset of respondents from the quantitative survey. Using purposive sampling, participants were selected to capture maximum variation in perspectives across specialities (Neurology, Geriatric Medicine, and Movement Disorders), levels of training (residents, early-career (<5 years of independent practice), and senior practitioners (>5 years of independent practice), and practice settings (independent, small, and tertiary care institutions). This approach ensured representation of physicians from diverse professional backgrounds across the country. Interviews were conducted till the saturation of themes was achieved—the qualitative phase aimed to explain and contextualize the quantitative findings.

Each interview was semi-structured, lasted approximately 30–45 minutes, and was conducted online via Zoom. Two trained medical students (PS and PK) conducted in-depth interviews (IDIs) using an interview guide (Supplementary appendix 1) developed from quantitative survey findings. The guide was designed to ensure the coverage of all major domains while allowing the exploration of emerging themes.

#### 2.2.2 Data collection

Both interviewers (PS and PK) underwent structured training in qualitative interviewing techniques, including formulating and asking open-ended questions, using neutral probes to encourage elaboration, and avoiding phrasing that could imply judgment or suggest that respondents were “wrong” for not engaging in ACP. The training aimed to minimize interviewer influence, reduce social desirability bias, and enhance consistency. They introduced themselves and informed the interview participants that they were final-year medical students. The interviews were conducted in English via Zoom (Zoom Video Communications Inc., San Jose, CA, USA), enabling remote data collection, interaction, and recording. The participants gave verbal consent to be recorded and were informed that they could end the interview at any time they felt appropriate. They could also revoke consent to use the interview data at any point, even after the interview was completed, provided that the manuscript had not been published.

### 2.3 Data analysis

#### 2.3.1 Quantitative study

We calculated the sample size required to estimate the item-level proportions of attitudes towards ACP, with a precision of 10%. Assuming that 50% of the respondents had knowledge of advance directives, 100 responses were required. Responses were collated in a spreadsheet using Microsoft Excel (Microsoft Office Excel 2019, Version 2407; Microsoft Corporation, WA, USA) and cross-verified for further analysis. The results for each question were analyzed individually based on the number of responses obtained.

The results were summarised using Stata version 17.0 (StataCorp LLC, College Station, TX, USA). The summary statistics are presented in numerical form, representing proportions as percentages. When more than one option could be selected as a response, each option was considered separately, and the proportions were calculated as either null or yes. We compared the respondents who participated in the qualitative study with those who did not. The data were reported in proportions, and between-group comparisons were performed using the chi-square test or Fisher’s exact test.

##### Qualitative Data Analysis

Interviews lasted 30–45 minutes and were audio-recorded, transcribed verbatim, and anonymized before analysis. The interviews were transcribed using Riverside (Riverside.fm, Tel Aviv, Israel), an AI-based transcription tool, and coded using NVivo 15 (NVivo software, QSR International, Melbourne, Australia) for thematic analysis. The transcription and coding were performed after each interview. Iterative coding was conducted, with initial codes developed both deductively from the CFIR framework and inductively from the data. After 2 or 3 interviews, the anonymized transcripts were reviewed by one author (AE). This was followed by reflexive periods before the next set of interviews, during which interviewers examined their own assumptions, reflected on potential bias, and adapted the interview process as needed. We defined thematic saturation as the non-emergence of new codes in further interviews.

## Results

### Quantitative Survey

A total of 140 physicians participated (Table 1). Of these, 44 (31.4%) were aged 20–30, 62 (44.3%) were 31–40, 17 (12.1%) 41–50, 12 (8.6%) 51–60, and 5 (3.6%) were above 60 years old. More than three-fifths of the respondents were affiliated with government medical colleges or teaching institutions. All respondents cared for patients with PD or other neurodegenerative disorders. Formal ACP training was reported by only 37.8% of participants, while approximately 15% had no exposure at all.

The attitudes and approaches of the respondents are summarised in Table 2. Attitudes toward ACP were largely positive. Most physicians (93.6%) agreed that ACP was necessary, and 51.4% preferred initiating discussions at the time of diagnosis. Most participants believed that the treating neurologists should lead the ACP conversations (54.3%). However, only 25.% reported sufficient time to conduct ACP discussions, and 65.7% felt they lacked adequate resources. While 74.3% felt mostly or always comfortable discussing ACP, 25.6% cited a lack of clarity on legal aspects, while others reported concerns about patient receptiveness and emotional distress. (Table 2)

### Qualitative Interviews

Of the surveyed respondents, 35.7% expressed willingness to participate in one-to-one interviews conducted as part of the qualitative component of the study. Based on demographic characteristics, 22 of them were contacted to ensure diverse representation by age, practice setting, geographic region, and level of training. Of the 22 contacted participants, seven declined participation due to time constraints or discomfort in discussing the topic. Ultimately, 15 participants were interviewed. Recruitment was continued until thematic saturation was achieved. These Qualitative interviews reinforced and expanded on the quantitative findings by revealing a range of themes. The themes developed from the qualitative analysis were organized using the Consolidated Framework for Implementation Research (CFIR)(13). These included intervention characteristics (perceived relative advantage, complexity, and adaptability), outer setting (patient needs and policy influence), inner setting (structural characteristics and implementation climate), individual characteristics (knowledge, training, and confidence), and process (engagement, execution, and reflection) (Table 3).

**Table 3.**
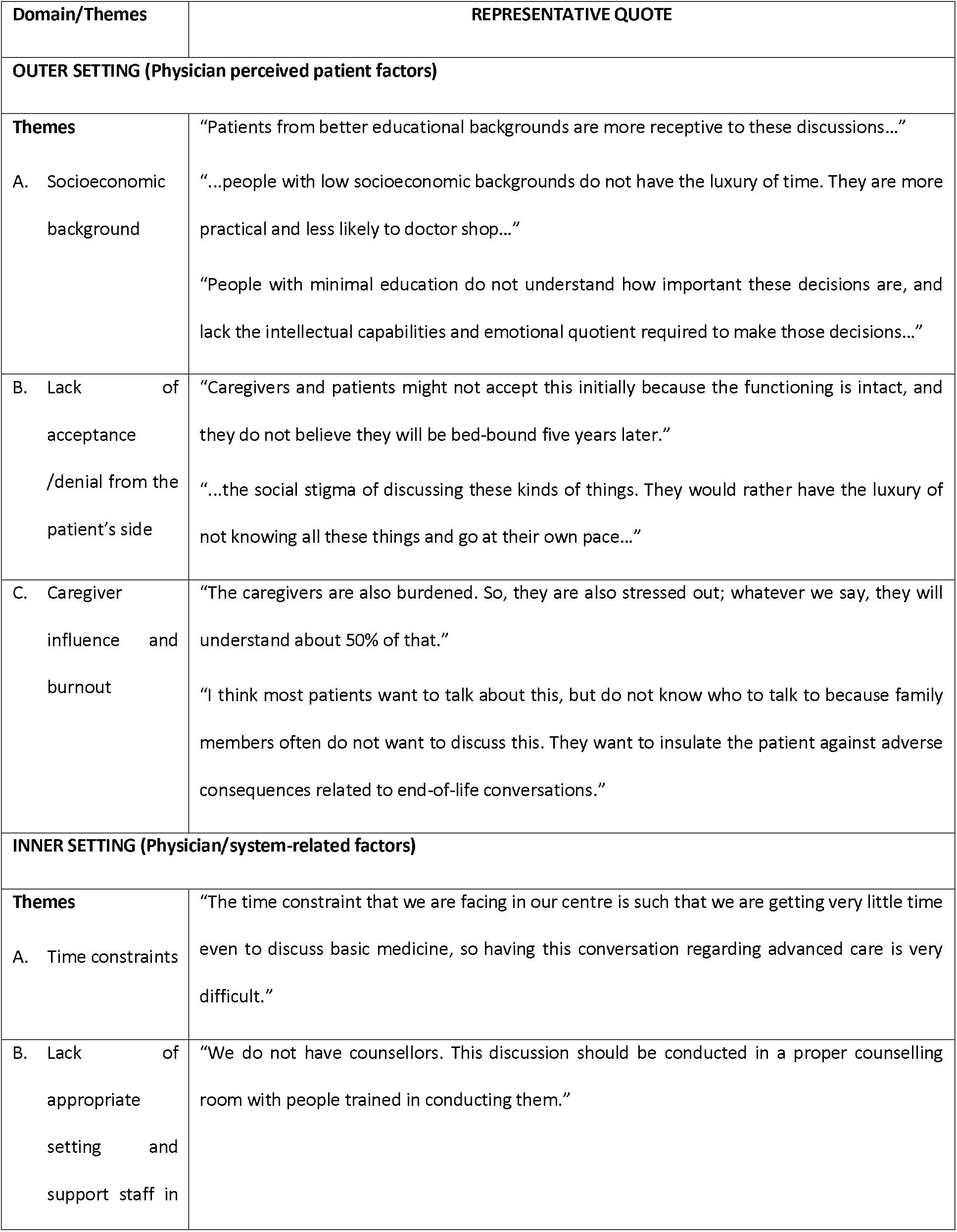
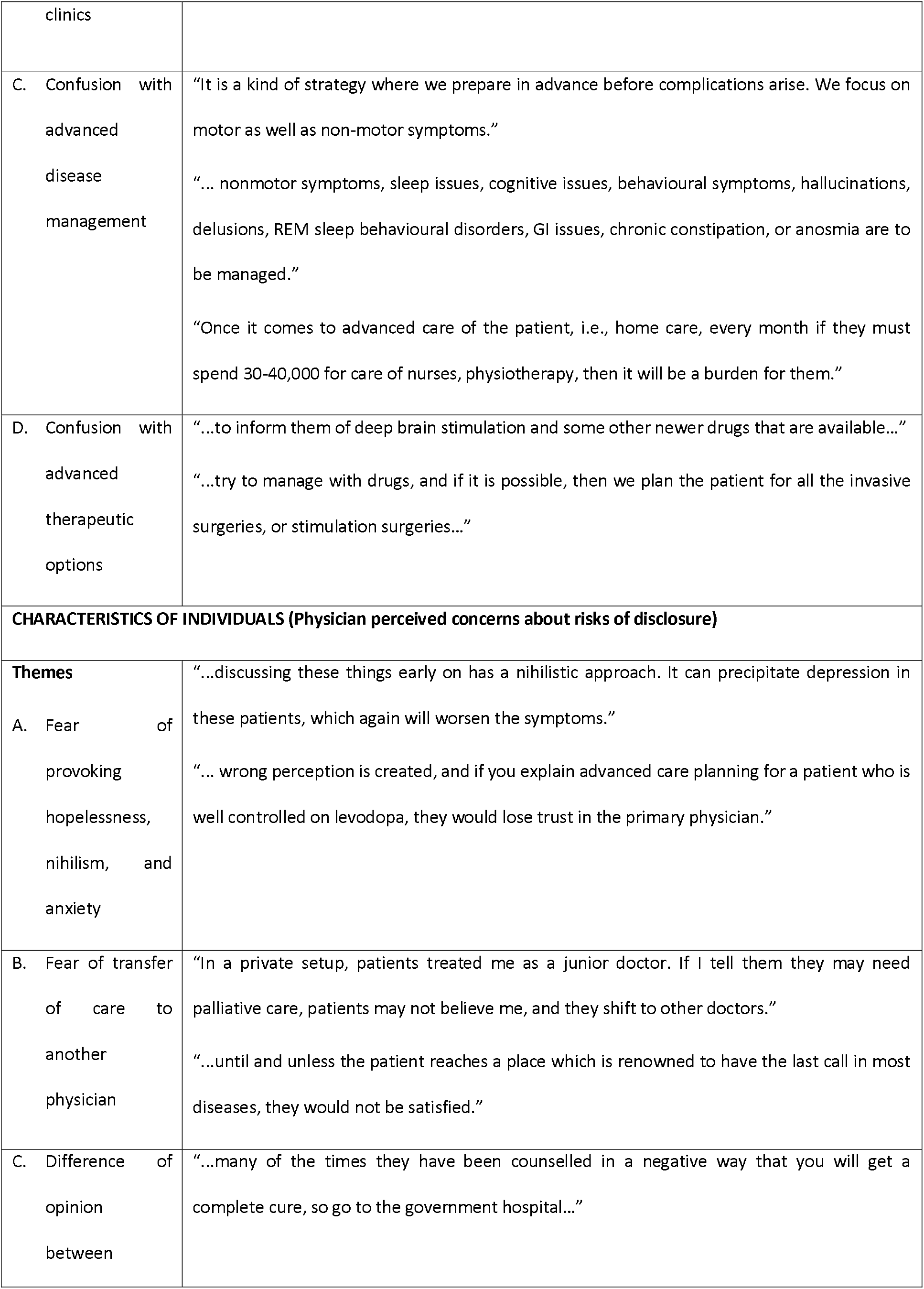
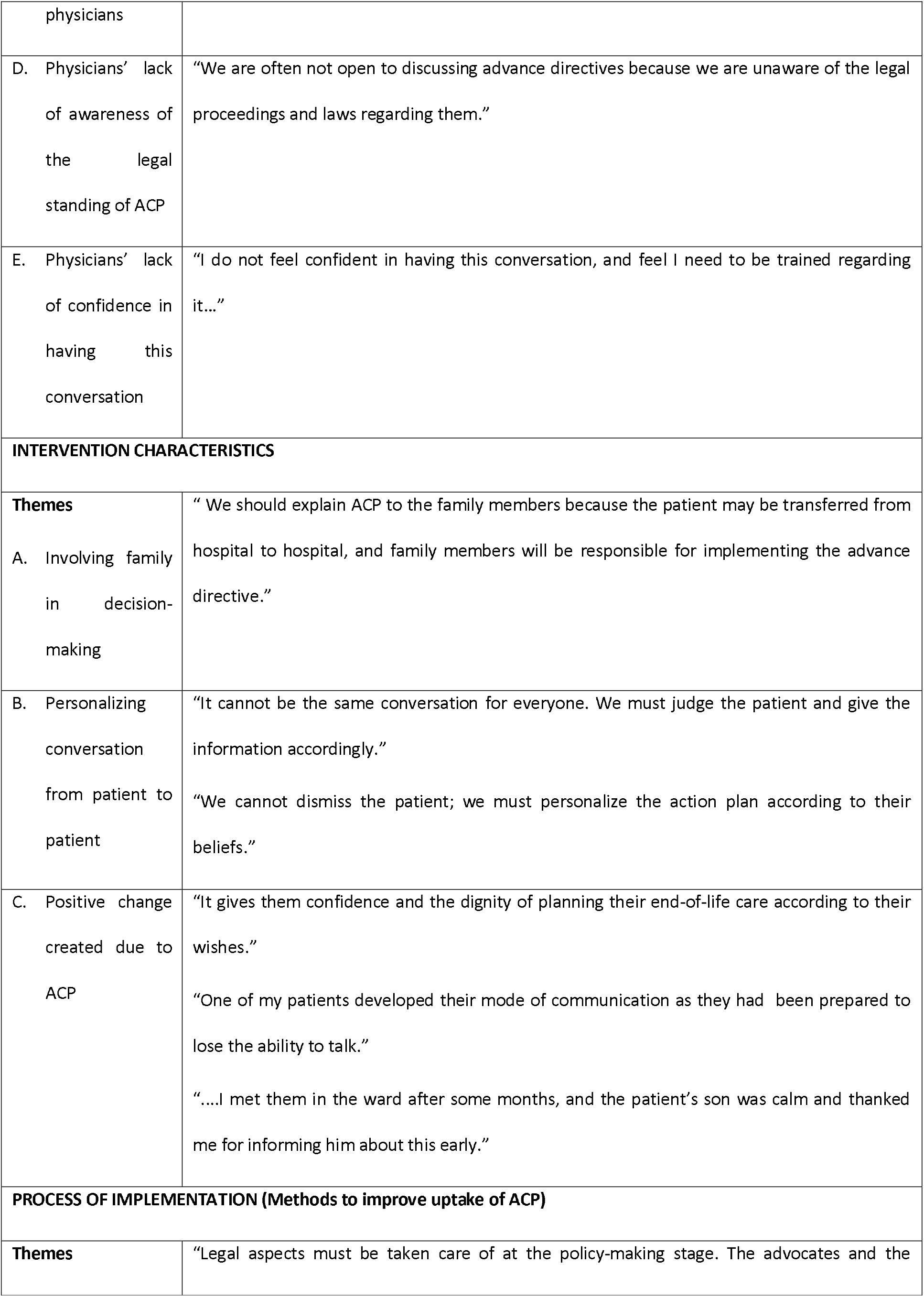
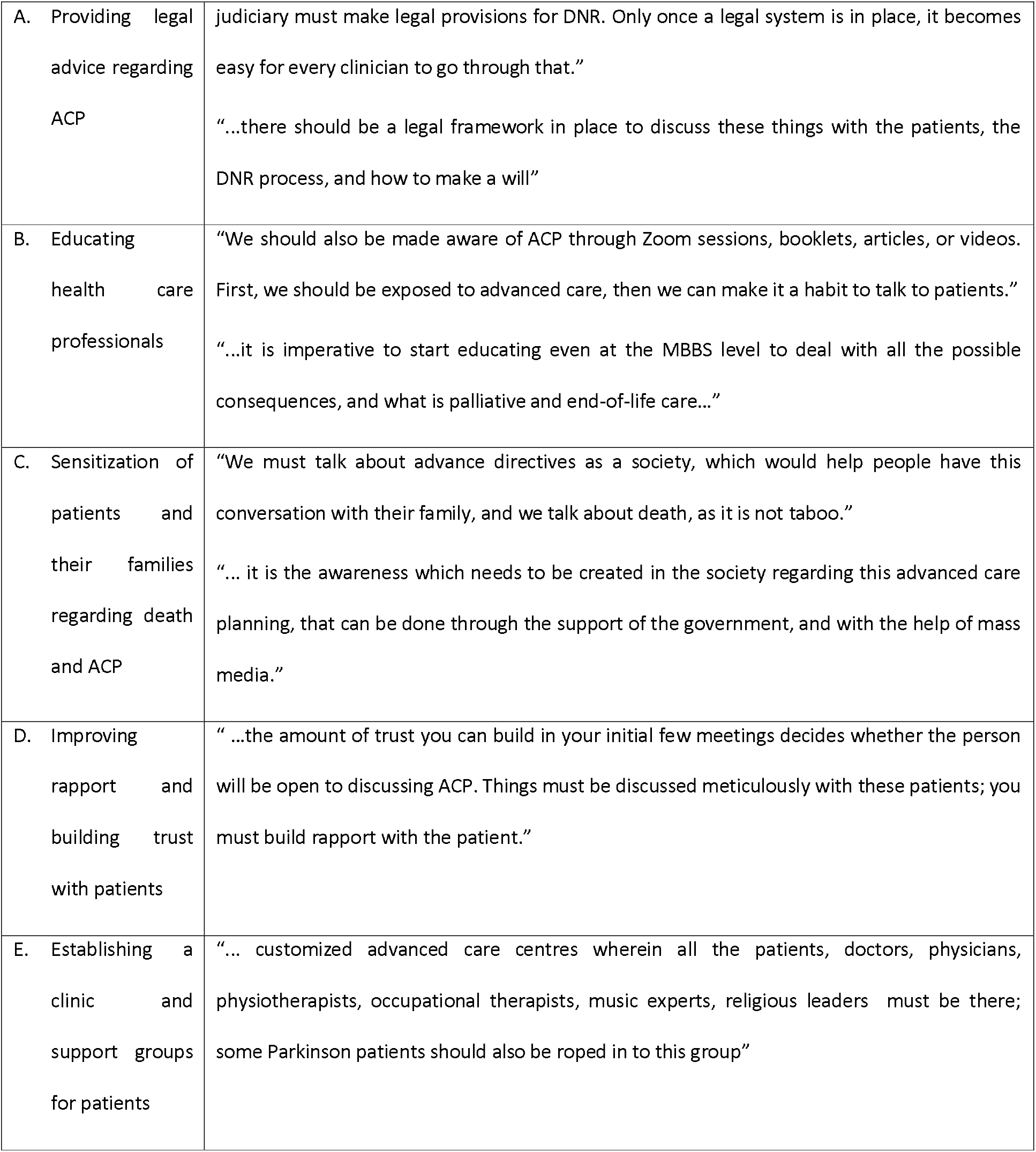
Themes that emerged from the Qualitative analysis described as per the Consolidated Framework for Implementation Research (CFIR) (13)

#### Theme 1: Physician perceived patient factors

Physicians frequently cited the patients’ socioeconomic and educational backgrounds as barriers to care. Patients with a limited education were perceived as less capable of understanding or engaging in ACP (Table 3). Denial of disease progression and discomfort when discussing death were thought to be common, especially among caregivers. The respondents felt that caregiver fatigue and resistance to emotionally charged conversations often prevented meaningful ACP engagement.

#### Theme 2: Physician perceived factors: Concern about risks of disclosure

Several physicians expressed concerns that initiating ACP discussions early on could provoke anxiety or diminish hope, especially in patients who have been recently diagnosed, are early in the course of the disease, or are still coming to terms with their illness. Some physicians also feared that these conversations might lead to a loss of trust or prompt patients to seek second opinions, particularly in private settings. Some participants highlighted inconsistencies in ACP practices across providers, which led to patient confusion and, consequently, resistance to engaging in these discussions (Table 3).

#### Theme 3: Physician/System-related factors

Time constraints were a significant issue, with physicians citing insufficient time during consultations for meaningful ACP. The lack of trained staff, counselling spaces, and institutional frameworks further hindered implementation. Many were unaware of the legal standing of ACP in India and lacked confidence in navigating such conversations. Confusion between ACP and advanced therapeutic options, such as deep brain stimulation and infusion therapies, also created communication challenges.

#### Theme 4: Current scenario of ACP

Despite these barriers, several physicians described positive outcomes when ACP was initiated early and tailored to the patient. Some noted that ACP enhanced trust and patient autonomy, with patients and families expressing gratitude. Involving families and personalizing discussions were considered essential strategies in the Indian context.

#### Theme 5: Methods to improve uptake of ACP

Participants recommended creating legal guidelines for ACP and DNR (do not resuscitate) orders, introducing ACP education in the medical curriculum from the undergraduate level, and conducting public awareness campaigns. They emphasized the importance of normalizing end-of-life discussions in clinical practice and broader societal contexts, highlighting that such conversations should not be treated as ‘taboo’. Multidisciplinary ACP clinics and support groups, involving clinicians, counsellors, and patients themselves (Table 3), were also proposed as a model for future and holistic care. A word cloud was generated using NVivo software to depict the most frequently recommended strategies as mentioned by the participants. (Figure 2)

**Figure 1.**
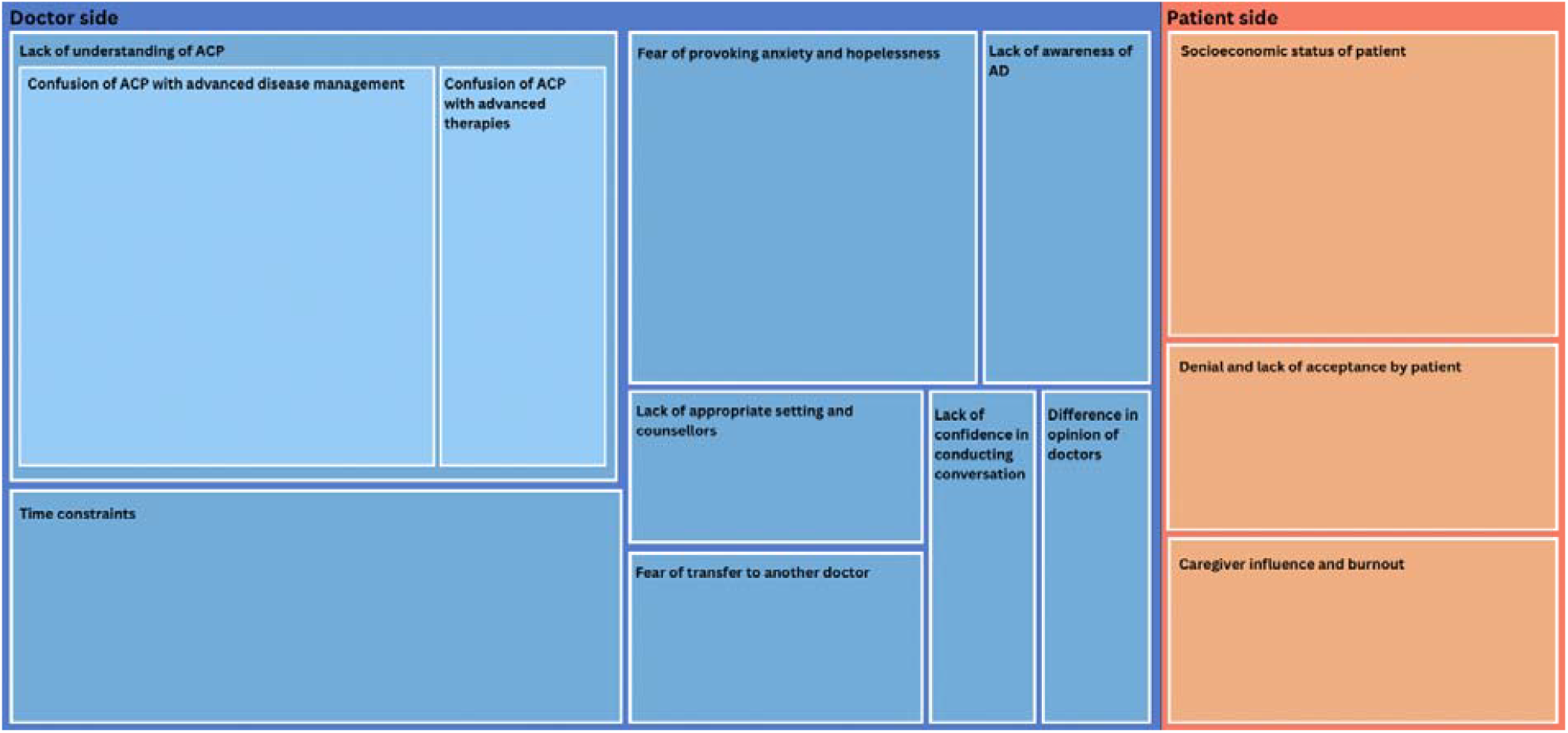
Hierarchy chart of various subthemes under the theme: Barriers to Advance Care Planning (ACP) The NVivo-generated visual layout hierarchy chart highlights the multidimensional challenges across stakeholders, as reported during in-depth interviews. Physician-level and system-level barriers are shown in blue, while physician-perceived patient-level barriers are represented in orange. The visual layout highlights multidimensional challenges across stakeholders, as reported in the in-depth interviews.

**Figure 2.**
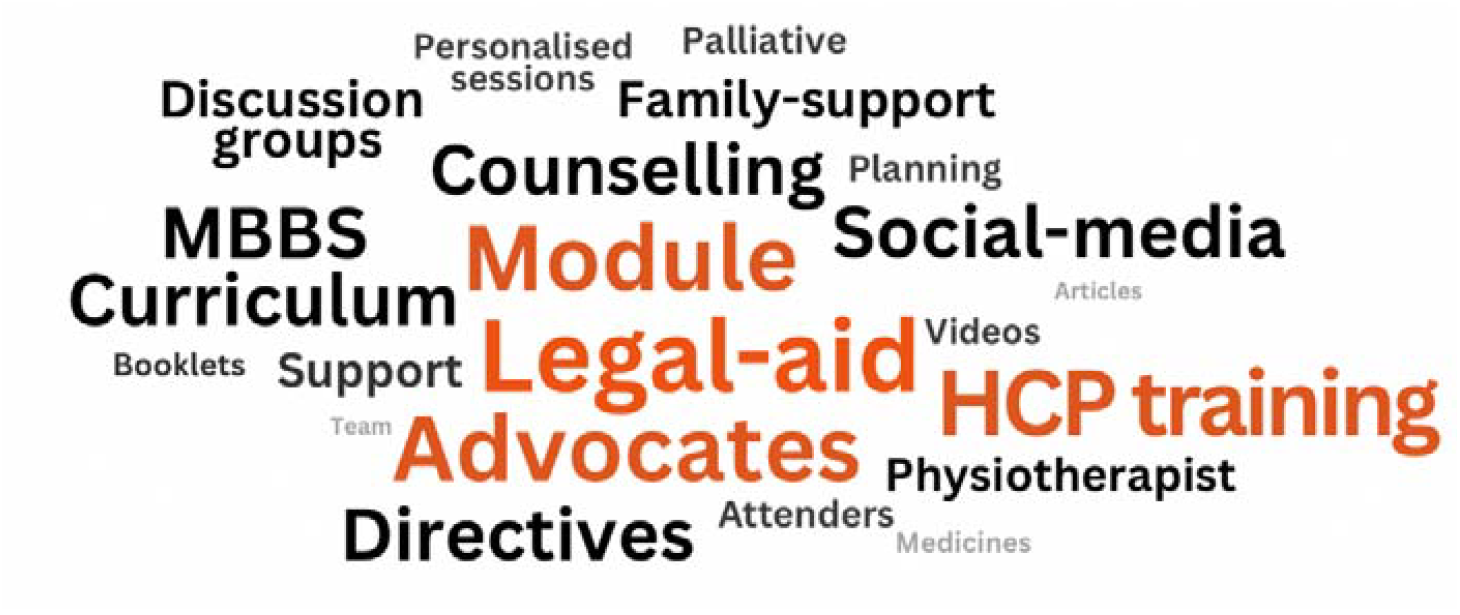
Word cloud representing the theme: Methods to Improve Uptake of Advance Care Planning (ACP) This word cloud was generated using NVivo software and reflected the physicians’ most frequently mentioned strategies during qualitative interviews. Terms such as “Legal-aid,” “Healthcare professional training,” “Module,” and “MBBS Curriculum” appear prominently, indicating their centrality in responses. Visualization captures key ideas for enhancing ACP engagement across clinical settings.

### Analytical Triangulation of Quantitative and Qualitative Data

A joint display (Figure 3) presents the integration of survey findings and interview themes, organized under CFIR domains. This analytical triangulation demonstrates convergence and complementarity between datasets, highlighting how systemic constraints, individual physician beliefs, and institutional challenges interact to influence the implementation of advance/future care planning.

**Figure 3.**
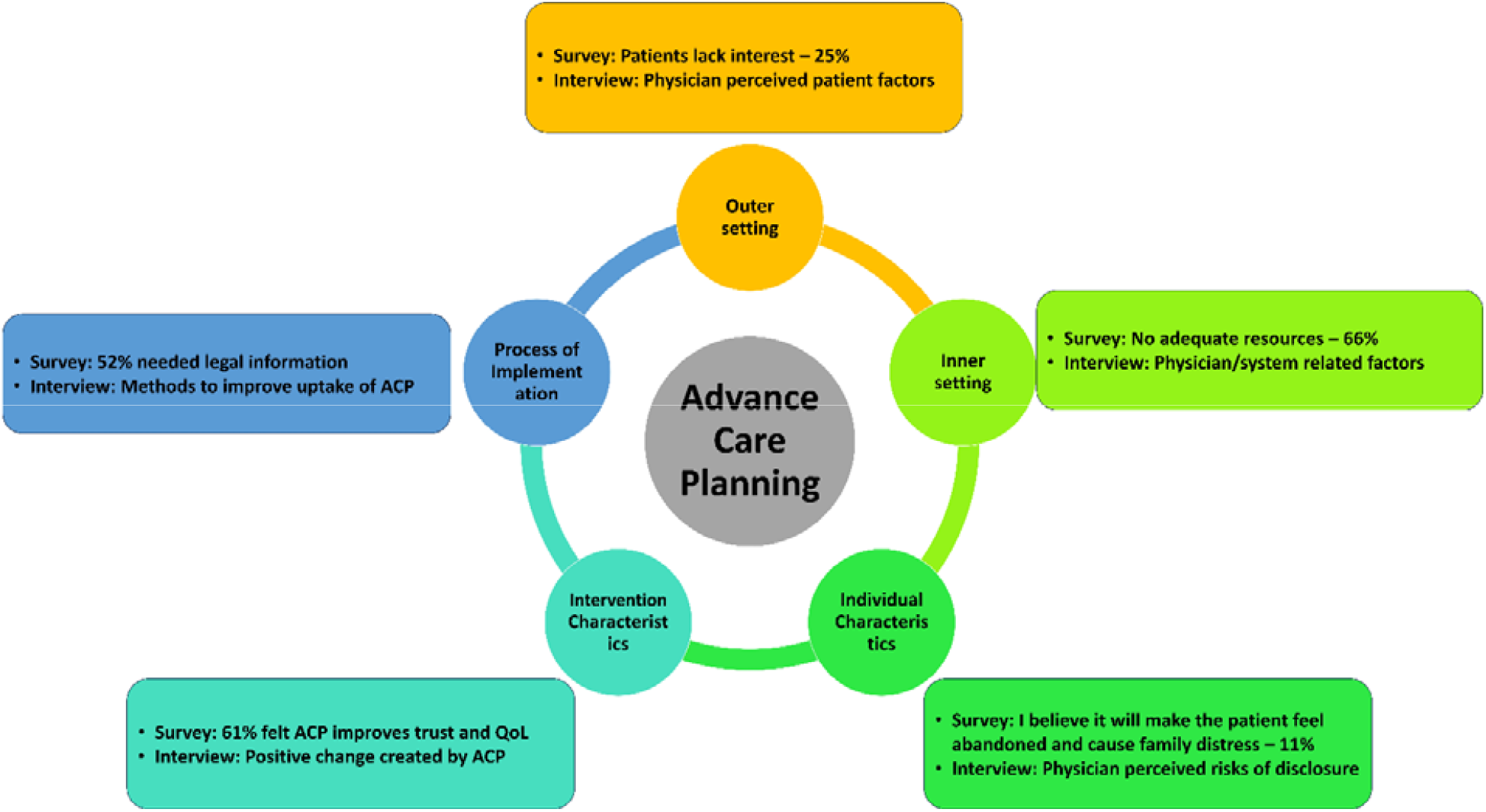
Triangulation of survey and interview data under CFIR domains, showing convergent and complementary insights into the implementation of advance/future care planning.

## Discussion

Our study explored physicians’ attitudes, practices, and barriers related to ACP in the context of Parkinson’s disease and other neurodegenerative disorders in India. We found that only a quarter reported having enough time for such discussions, and 65.7% indicated inadequate resources, including a lack of appropriate space for discussion and support from counsellors. Additionally, many physicians in our study reported lacking the training and confidence to conduct such sensitive conversations. Several physicians seemed to have a lack of understanding regarding the meaning of advance care planning and confused it with advanced Parkinson’s disease and advanced therapies. This trend has also been observed in other low-to middle-income countries, such as Peru, where a study revealed that only a quarter of doctors knew what an Advanced Medical Directive(AMD) was. They reported discomfort in conducting these conversations and a lack of legal clarity regarding documentation (14). Despite the 2018 Supreme Court judgment legalizing living wills (15), confusion persists around their implementation, documentation, and legal validity, making physicians hesitant in having conversations regarding ACP.

Professional apprehensions also emerged prominently. Physicians were concerned that ACP discussions might be misinterpreted as pessimism or a loss of hope, potentially leading to a breakdown in trust or patient attrition, particularly in private practice settings. However, physicians from Western countries are less likely to withhold unfavourable information from the patient at the family’s request, avoid discussing the issue entirely, use euphemisms, or administer treatments known to be ineffective in order not to destroy hope (16). A study conducted in Canada on a population of Southeast Asian communities found that sociocultural barriers prevented ACP (17).

In our study, physicians perceived that patients would not accept ACP due to denial of disease, limited education, lack of understanding, and the influence of family. Indian society often adopts a collective and family-centred approach to care, where decision-making is deferred to family members, and discussions around death are considered inauspicious, as seen in similar studies from Asian countries (18,19). While protective in some ways, these cultural norms may hinder honest conversations about future medical care and end-of-life preferences. In a study conducted in five Canadian provinces by You et al., clinicians perceived their skills and system factors as less significant barriers (20). In contrast, our findings revealed these to be significant factors (Figure 1). This could also be because of the inherent biases of the Indian Physicians regarding patient and caregiver perspectives.

Several physicians shared positive experiences when ACP was implemented early and personalized to the patient’s values and cultural context. This supports findings from Western and Asian settings where culturally appropriate and family-inclusive ACP enhances patient autonomy, improves end-of-life care, and strengthens trust in the healthcare provider(21,22). Family involvement was viewed as essential in our study, consistent with collectivist cultures where family plays a central role in healthcare decision-making. (23).

This study has several strengths. It employed a mixed-methods approach, which enabled both a systematic assessment of attitudes and practices in a larger cohort and rich contextual insights into ACP in a subset. The comprehensive questionnaire captured diverse perspectives from physicians, and the anonymous, self-administered format helped reduce social desirability bias. Importantly, the interviews were conducted by medical students, which further minimized hierarchical influences compared to those of senior colleagues or administrators, likely encouraging openness and strengthening the authenticity of the qualitative data. Having medical students conduct the research process also strengthened the study, as the interview was guided by individuals who are less conditioned by established local clinical practices.

This study had several limitations. First, the study was subject to selection bias owing to its online recruitment, small sample size, lack of random sampling, and self-selection bias, which are common issues in most online surveys. The study was not powered to detect subgroup differences between the qualitative and quantitative parts. As the sample focused on physicians from teaching hospitals, it may not reflect all practice settings. Self-reporting and willingness to participate may lead to selecting respondents who are already interested in ACP. We were unable to determine the actual reach of the survey as it was disseminated through social media. This may limit the generalizability of the findings to diverse healthcare settings nationwide. The absence of an interviewer for the survey portion meant we could not verify participants’ understanding of the questions, as evidenced by their responses during the qualitative interview. Several participants misunderstood ACP, specifically regarding treatment for advanced disease stages and advanced therapeutic options such as deep-brain stimulation. Although ACP was defined and briefly described in the consent form at the beginning of the survey and during the interview, some participants still held misconceptions, which were non-quantifiable in extent. Nonetheless, online surveys are increasingly recognized for their efficiency and accessibility, particularly in professional communities. We did not study the perspectives of practitioners of alternative medicine, nurses, palliative care workers, or other allied health professionals.

Additionally, as a cross-sectional survey, the data reflect attitudes and practices at the time of data collection and do not capture changes over time owing to the impact of institutional or policy shifts. While measures were taken to limit social desirability bias, it may still have influenced the responses, particularly in self-reported practices related to sensitive topics, such as end-of-life care. Finally, although the qualitative themes provided deeper insights, the open-ended responses varied in detail and depth.

Based on the insights gained from this study, we propose strategies to enhance the implementation of ACP in India. First, there is a need for national legal and ethical guidelines that clearly define ACP processes, documentation, and the responsibilities of physicians. The 2023 Supreme Court of India amendment streamlined the legal framework for AMD by removing the requirement for judicial oversight and enabling execution before a notary or a gazetted officer, thereby facilitating greater accessibility. Although it has increased physicians’ confidence in implementing ACP, there is an urgent need to develop modules for physicians to make advance care planning a more comfortable discussion topic. There is a need to promote death literacy, which refers to the practical knowledge and skills that enable individuals and communities to understand, plan, and make informed decisions about death, dying, and end-of-life care, and increase awareness of available care options, legal rights, and communication around dying, to help people navigate these experiences with greater confidence and compassion. There is also a need to explore religious, spiritual, and cultural aspects so that they can be incorporated into the medical curriculum, ensuring a sensitive and culturally appropriate approach to ACP. This will enable clinicians to initiate and navigate conversations more effectively. ACP should also be incorporated into the National Program for Healthcare of the Elderly (NPHCE) and National Program for Palliative Care (NPPC), run by the Ministry of Health and Family Welfare(MoHFW), and in the comprehensive care package delivered from Arogya Mandir (AAM) centres under the Ayushman Bharat program. Because patients in India often seek advice from several experts, discrepancies in prognostic assessments are common, underscoring the need for uniform evidence-based guidelines. Establishing multidisciplinary clinics and support services, comprising physicians, palliative care specialists, counsellors, and social workers, could provide the structural and emotional scaffolding needed for meaningful ACP conversations. There is also a need to understand patient and caregiver perspectives on ACP, the appropriate time to initiate ACP for individual diseases, and assess changes in trends over time, particularly in response to evolving legal, cultural, and educational initiatives.

The use of the term ‘advance care planning’ in neurodegenerative disorders may be problematic, as it can be misinterpreted as treatment of advanced disease - or end-of-life care–thereby limiting its acceptance in practice. The availability of advanced therapeutic options used in various phases of these illnesses also contributes to the confusion. Replacing it with ‘Future care planning’ could provide greater clarity and prevent confusion with advanced treatment modalities or the management of Advanced stages of Neurodegenerative diseases.

## Conclusion

This study provides valuable insights into the attitudes, practices, and perceived barriers to advance care planning among physicians caring for patients with Parkinson’s disease and other neurodegenerative disorders in India. While most physicians acknowledged the necessity of ACP, its implementation in clinical practice remains limited due to barriers. Despite these barriers, physicians were willing to engage in such conversations, particularly when appropriate frameworks and support systems are in place. One of the key strengths of this study is its mixed-methods approach, which combines quantitative data with qualitative insights, enabling a deeper understanding of both the measurable and contextual challenges faced by physicians. These findings underscore the urgent need for structured ACP training in medical curricula and institutional policies that enable clinicians to meaningfully engage patients and families in end-of-life planning.

## Data Availability

The datasets generated and/or analyzed during the current study are not publicly available due to the inclusion of identifiable qualitative data but are available from the corresponding author on reasonable request. De-identified quantitative data may be shared upon request for academic and research purposes.

## Ethical Compliance

The protocol was approved in the meeting of the Ethics Committee for Post Graduate research (Basic science) of the All India Institute of Medical Science, New Delhi, held on 26.09.2024.

## DECLARATIONS

### Ethics approval and consent to participate

The study was approved by the Institute Ethics Committee for Post Graduate Research, of the All India Institute of Medical Sciences, Ansari Nagar, New Delhi, in the meeting conducted on 26^th^ September, 2024, and the protocol was approved from the ethical angle.

Consent: All participants provided informed written consent to participate in the study. For the qualitative interviews, informed verbal consent was video recorded from all participants prior to data collection, ensuring confidentiality and voluntary participation.

### Consent for publication

All authors have read and approved the final version of the manuscript and consent to its publication in the *International Journal for Equity in Health*.

### Competing interests

The authors declare that they have no competing interests.

### Funding

This study received no specific grant from any funding agency in the public, commercial, or not-for-profit sectors.

### Authors’ contributions

Pavit Singh – Study design and implementation

Parvathy – Study design and implementation

Deepa Dash – Conceptualization, critique of manuscript

Nishkarsh Gupta – Conduct of study

Usha Ramanathan – Critique of manuscript

Teneille E. Gofton – Critique of manuscript

Claudia Chou – Critique of manuscript

Soaham Desai – Critique of manuscript

Pramod Kumar Pal – Critique of manuscript

Roopkumar Gursahani – Critique of manuscript

Baikuntha Panigrahi – Critique of manuscript

Banusri Velpandian – Critique of manuscript

Prasun Chatterjee – Critique of manuscript

Avinash Chakrawarty – Critique of manuscript

Anup Singh – Critique of manuscript

Suman Kushwaha – Critique of manuscript

Divya K.P. – Conduct of study, critique of manuscript

Lakshmi Narasimhan Ranganathan – Conduct of study

Manjari Tripathi – Conduct of study

Deepti Vibha – Conduct of study

Rajesh Kumar Singh – Conduct of study

Animesh Das – Conduct of study

Jasmine Parihar – Conduct of study

Sumit Malhotra – Conduct of study

Ashish Dutt Upadhyay – Statistical analysis

Abhishek Pathak – Critique of manuscript

Arunmozhimaran Elavarasi – Conceptualization, conduct of study, statistical analysis, review, and overall responsibility for the study

## Acknowledgements

Anup Singh (AS) would like to acknowledge the National Programme for Health Care of the Elderly (NPHCE), MoHFW, Government of India, for support.

## Notes

### Competing Interest Statement

The authors have declared no competing interest.

### Funding Statement

No funding received

### Author Declarations

The study was approved by the Institute Ethics Committee for Post Graduate Research, of the All India Institute of Medical Sciences, Ansari Nagar, New Delhi, in the meeting conducted on 26th September, 2024, and the protocol was approved from the ethical angle.

