## Supplementary appendix for "Barriers and Facilitators to Advance Care Planning Implementation for Patients with Neurodegenerative Diseases among Indian Physicians: A Mixed-Methods Analysis"

**In-Depth Interview Guide: Neurologists and Advance Care Planning in Parkinson's & Neurodegenerative Diseases**

A guide for the primary investigators who are conducting the qualitative study

### Opening/Rapport building

- Introduce yourselves, and greet the participant
- Show slide, explaining the study and the definition of Advance care planning.

#### Explain purpose

- *"We are trying to understand how clinicians think and feel about Advance Care Planning (ACP) and palliative care in the context of Parkinson's and related disorders — especially the emotional, systemic, and practical aspects that are not always captured in surveys.* *There are no right or wrong answers; we are interested in your perspective "*
- *Approved by the Institute Ethics Committee*

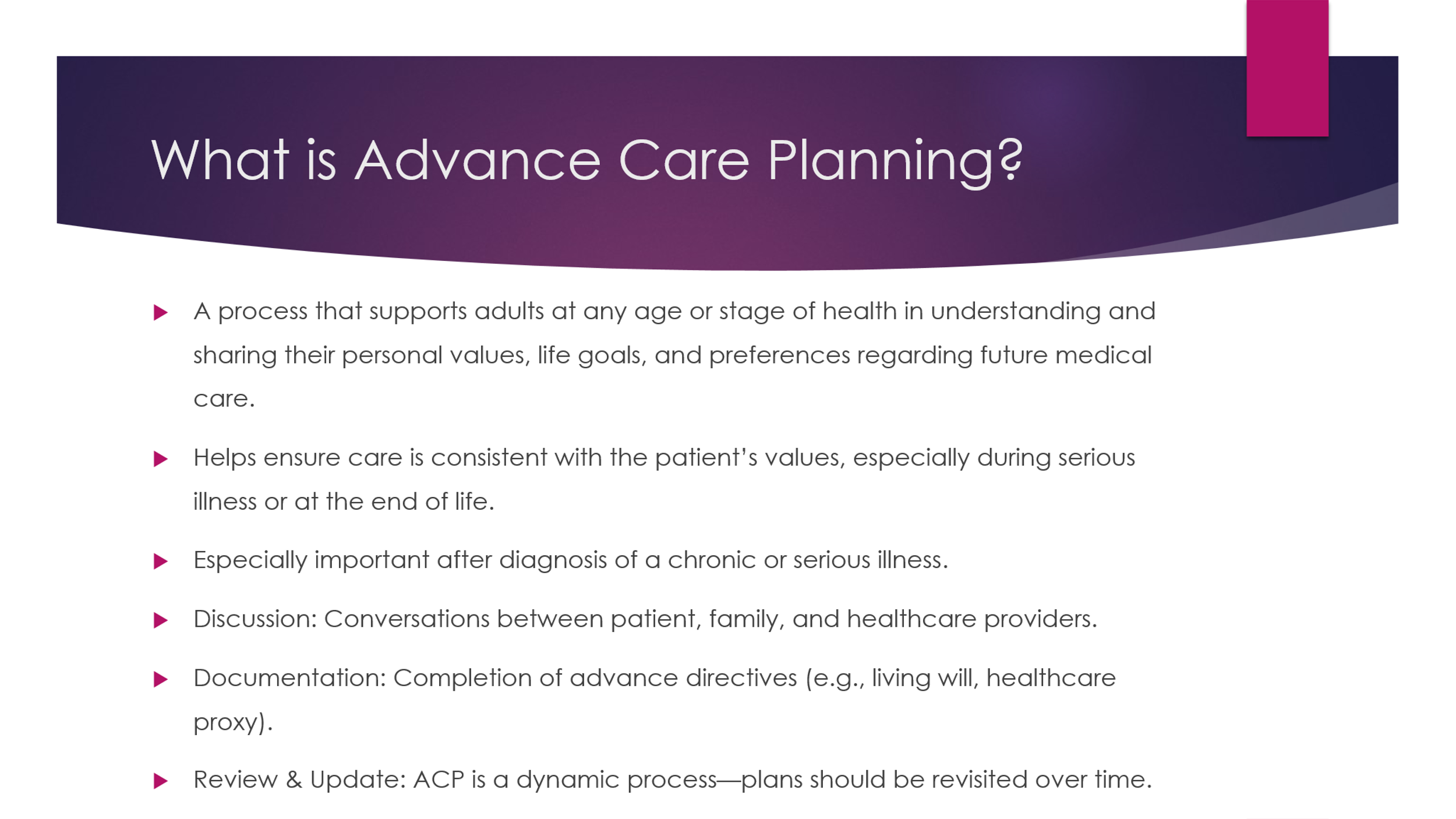

- This slide should be on the screen during the entire interview process

#### Take Verbal consent

- To use information and quotations in the publication
- Permission to record the Zoom call and then switch on the recording

#### Assure confidentiality

### Introduction to the Topic

**What are your general thoughts on Advance care planning**

### Key domains/Themes

Try to cover these, but avoid putting too much pressure on yourself to complete all questions; let the conversation flow naturally.

1. **What impact do you believe ACP will have on a patient with Parkinson's disease?**
2. **What have you observed about the perspective of patients who accepted vs rejected ACP?**
3. **How do you personally recognize that it is time to start conversations around ACP in a patient's journey?** *→ clinical cues, patient-family signals, personal experience*
4. **Can you share a case that shaped the way you now approach ACP in practice?** *→ experience-driven insights*
5. **In your experience, what are the most significant barriers to implementing ACP effectively in your setting?** *→ Time, resources, family resistance, patient readiness, etc.*
6. **What kind of support (systems, training, personnel) do you think would make it easier to integrate ACP into routine care?**
7. **Have you ever felt that a discussion about ACP changed the relationship between you and a patient/family?** *→ successful communication models, soft skills, or trust-building techniques.*
8. **What would make you feel more confident or better equipped to have these conversations regularly?** *→ legal clarity, scripts, case-based training, supervision, etc.*
9. **Would you like to provide any additional comments or offer further insight regarding ACP?**

#### Clarification and Reflection

Ask the participant to reflect or summarize the key views

#### Closing

- Thank the participant, ask if they want to add anything
- Reassure confidentiality, tell them they can call back any time before the paper is published to withdraw consent to participation

### Tips and tricks

- Aim for 35-45 minutes
- Do not ask confronting questions
- Do not ask leading questions
- Do not ask yes/no questions
- Let the participant speak, do not interrupt
- If some point is not clear, ask them to repeat – do not paraphrase and ask them if what you said is correct or not.
- Do not make the participant feel that what they are doing or practising is wrong – there are no right or wrong answers here. Try to understand their perspective and document what they are saying. This exercise is to understand their views. Not to teach them about ACP or how to implement it.
- After the interview, transcribe the audio as soon as possible and start coding.
- Do not do more than two interviews in a day.
- Reflect on the interview, read the transcript, and make sure your biases are not creeping into the interview process.
